# AI-supported automated microscopy for malaria diagnosis

**DOI:** 10.1101/2025.06.11.25329408

**Authors:** Dawit Hawaria, Yalemwork Ewnetu, Henry Kamugisha, Abdul-Hakim Mutala, Birhanu Lulu, Colins O. Oduma, Isaac Asenso Brobbey-Kyei, Tolulope Adeyemi Kayode, Wossenseged Lemma, Nega Berhane, Kingsley Badu, Cristian Koepfli

## Abstract

**Background:** Accurate malaria diagnosis is key for patient management, surveillance, and control. Automated microscopy can overcome variation observed among microscopists and is a promising new tool for diagnosis. The Noul miLab integrates smear preparation, staining, imaging, and AI-supported parasite detection in a portable device.

**Methods:** 2201 samples were collected from febrile patients across two sites in Ethiopia, where *P. falciparum* and *P. vivax* are frequent, and Ghana, where *P. falciparum* transmission is intense. Samples were screened by local microscopy at the health center, miLab, and rapid diagnostic test. qPCR and expert microscopy were used as gold standard.

**Results:** Across the three sites in Ehtiopia and Ghana, miLab reached a sensitivity for *P. falciparum* diagnosis of 96.3% (335/348) using expert microscopy as gold standard, and of 97.4% (298/306) using qPCR-positive infections at densities >200 parasites/µL as gold standard. Across two sites in Ethiopia, the sensitivity of miLab for *P. vivax* was 96.8% (399/412) using expert microscopy as gold standard, and 95.9% (419/437) using qPCR-positive infections at densities >200 parasites/µL as gold standard. Specificity compared to qPCR was 98.8% (1057/1070) for *P. falciparum* and 97.8% (617/631) for *P. vivax*. The miLab was significantly more sensitive than microscopy conducted at the health center. In Ethiopia, the miLab assigned the correct species to 99.3% (147/148) *P. falciparum* and 96.5% (304/315) *P. vivax* mono-infections infections where a species was determined.

**Conclusions:** The miLab automated microscope shows high sensitivity and specificity for *P. falciparum* and *P. vivax* diagnosis.

## Background

Malaria is a major global health threat. In 2023, nearly 600,000 deaths and over 260 million clinical cases were reported [1]. While the main burden is in sub-Saharan Africa, malaria remains prevalent in many countries in Asia, Latin America, and the Pacific. Accurate and rapid diagnosis of febrile patients at health centers is a key pillar of control. The WHO recommends testing of all suspected cases of malaria. Antimalarial treatment should be limited to patients with positive tests [2]. For over 100 years, light microscopy had been the main tool for diagnosis. The quality of light microscopy depends heavily on the training and skill of the microscopists. As a result, a substantially lower number of infections is detected by routine microscopy conducted at health centers compared to highly trained expert microscopists [3, 4]. Where control is successful and the number of cases decreases, the sensitivity of microscopists has been shown to decline, resulting in more infections that are missed [5, 6].

In the last two decades, rapid diagnostic tests (RDTs) became a main alternative to light microscopy [7]. While the sensitivity of RDTs increased greatly over time, concerns remain about false-negative and false-positive results. The most common RDTs for *P. falciparum* detect the HRP2 and HRP3 antigens. Deletions of *hrp2* and *hrp3* genes result in false-negative tests. Deletions are frequent in the Horn of Africa, e.g., Ethiopia, and in Latin America [8]. RDTs might yield false-positive results as antigens can persist for up to several weeks after treatment is administered [9–11]. Trust in RDTs can further be undermined by false-negative results due to prozone effect (very high antibody titers in the host blocking the detection by RDT) [12], or due to potential manufacturing or storage issues [13].

Microscopy remains widely used for malaria diagnosis in health centers and hospitals. The direct ability to see the parasite on a blood slide (compared to only a test line on an RDT) increases trust in diagnosis by doctors and patients. In addition to showing the presence or absence of parasites, microscopy yields data on parasite density, and stages of the parasite present. To overcome the variation in quality of microscopists, automated microscopy tools have been developed [14]. Some tools include smartphone-based apps that analyze images [15, 16]. They still require manual preparation and staining of slides. Other systems incorporate all steps from preparing a smear to staining, reading, and data interpretation [17]. In its ‘Global Malaria Programme operational strategy 2024-2030’, the WHO listed further development of automated microscopy as a key component for diagnosis [18].

The miLab, developed by Noul Inc, is the first commercially available fully integrated AI-supported automated microscope for the diagnosis of clinical malaria cases. In a previous study, we showed robust performance for the diagnosis of *P. falciparum* and *P. vivax* [19]. Here, using an updated algorithm, we screened 2201 samples in Ethiopia and Ghana and assessed the performance of the miLab using expert microscopy and qPCR as reference. In particular, we determined the ability to distinguish between *P. falciparum* and *P. vivax* among 1596 samples collected in Ethiopia where the two species are co-endemic.

## Methods

### Ethical Approval

Informed written consent was collected from each adult, or, in the case of minors, from the legal guardian prior to sample collection. This study was approved by the Hawassa University College of Medicine and Health Sciences Institution Review Board (ref. no. IRB/355/15), the University of Gondar Vice President for Research and Technology Transfer (Ref VP/RTT/05/546/2024), the Kwame Nkrumah University of Science and Technology Committee on Human Research, Publication, and Ethics, College of Health Sciences (ref: CHRPE/AP/952/23), and the University of Notre Dame Institutional Review Board (approval no. 23-07-7990).

### Diagnostic tests

The intended use of the Noul miLab automated microscope (the index test) is the automated microscopic examination of whole blood samples for the detection of *P. falciparum* and *P. vivax* infections among febrile patients at health centers and hospitals. The miLab is intended to be used by professional users, trained in the use of the device. The operator loads 5 µL of blood into a cartridge and inserts it into the miLab, which prepares a smear, stains it, and detects parasites using an AI-algorithm. Time to result is approximately 15 minutes. The miLab detects the number of infected red blood cells (RBCs) among approximately 200,000 RBCs. It displays results either as ‘suspected *P. falciparum*’, ‘suspected *P. vivax*’, ‘suspected *Plasmodium*’, ‘Negative’, or ‘Review needed’. For the current study, results displayed as ‘Review needed’ were counted as negatives. If parasites are detected, they are displayed on the screen of the device. Each infected RBC can be selected, and 10 images along the Z-axis can be displayed. The operator can thus verify the result of the algorithm.

As a comparator, samples were screened by RDT. In Ethiopia and Ghana, samples were screened by the BIOCREDIT Malaria Ag Pf (pLDH/HRP2) (Rapigen, South Korea). This RDT has separate test lines for HRP2 and *P. falciparum*-specific LDH. It thus allows detection of *P. falciparum* infections with *hrp2*/*3* deletion through the LDH line. In Ethiopia, samples were also screened with the BIOCREDIT Malaria Ag Pf/Pv (pLDH/pLDH) (Rapigen, South Korea). This RDT has separate lines for *P. falciparum* and *P. vivax*-specific LDH. Further, the result of the routine health center diagnosis conducted by microscopy was recorded. qPCR and expert microscopy were conducted as reference standards.

### Study sites and sample collection

Samples were collected in two sites in Ethiopia, Hawassa in the south and Gondar in the north of the country. In these sites transmission of *P. falciparum* and *P. vivax* is intense. Further, the frequency of *P. falciparum hrp2*/*3* deletion is high, in particular in Gondar [20]. In Hawassa, Ethiopia, samples were collected in three health centers (Alamura, Millenium, Tilte) between September 12, and December 25, 2024. In Gondar, Ethiopia, samples were collected in one health center (Maraki) between January 16 and February 26, 2025. In Ghana, samples were collected in and around Kumasi in the Ashanti region where *P. falciparum* transmission is intense. Samples were collected in three health centers (Agona Government Hospital, Asokwa Children Hospital, and Ejisu Government Hospital) between November 4 and December 20, 2024.

In all sites, individuals above 1 year of age presenting for malaria diagnosis were eligible to be enrolled. Patients were enrolled prospectively, and consecutively. After informed consent was obtained, from each patient, 200-250 μL of capillary blood was collected into microtainer EDTA tubes by finger prick. On site, samples were screened by miLab using software version 1.2 and by RDT, a slide for expert microscopy was prepared, and the result from the health center microscopist was recorded. The remaining blood was stored at −20 °C until DNA extraction. Operators of the index test (miLab) were not blinded to results by RDT and local microscopy, but did not have access to results by qPCR and expert microscopy which were conducted later.

### Expert microscopy and qPCR

For expert microscopy, 3 μL of thick and 2 μL thin blood smears were prepared and stained according to the WHO standard [7]. Slides from Hawassa and Gondar were read in Ethiopia, while slides from Ghana were shipped to the Kenyan Medical Research Institute (KEMRI) in Kisumu, Kenya, and read there. In either site, slides were read by two level 1 microscopists that were blinded to each other, and a third level 1 microscopist in case of discrepant results. A slide was declared negative after examining 100 microscopic fields. Asexual parasite density was calculated whenever a slide was reported as positive. Expert microscopists were blinded to the results of the index test.

For samples from Hawassa and Gondar, DNA extraction and qPCR were conducted at Hawassa University. Samples from Ghana were shipped on ice to the University of Notre Dame. Identical protocols were used in both sites. DNA was extracted from 100 μL blood using the Genomic DNA Extraction kit (Macherey-Nagel) and eluted in an equivalent volume of elution buffer. 4 μL of DNA, corresponding to 4 μL of blood, was screened for *P. falciparum* and *P. vivax* separately either on the MIC (Bio Molecular Systems) or QuantStudio 3 (Thermo Fisher) qPCR instruments. *P. falciparum* was detected using the varATS assay which amplifies a target present in approximately 20 copies per parasite [21], and *P. vivax* the *cox1* assay [22], which amplifies a mitochondrial gene present in approximately 10 copies per parasite. The 95% limit of detection is approximately 0.3 parasites/μL blood for *P. falciparum*, and slightly higher for *P. vivax* [23]. Absolute parasite densities for *P. falciparum* and *P. vivax* were obtained using an external standard curve quantified by droplet digital PCR (ddPCR). In the case of *P. vivax*, a higher density blood sample identified by microscopy was used to construct the standard curve, and for *P. falciparum*, 3D7 cultured parasites were used. Scientists conducting qPCR were not blinded to the results of the index test.

### Data analysis

All data analysis was done in Stata version 16.1 or R. Sensitivity of the miLab (the index test) was calculated separately using expert microscopy and qPCR as gold standards. For qPCR, the sensitivity was calculated against density thresholds of 20 and 200 parasites/µL. This was done to account for the fact that qPCR detects very low-density infections that are not expected to be detected by miLab or any other point-of-care diagnostic device. miLab data was transformed into parasites per µL blood as follows, assuming 5 million RBCs per µL: (5,000,000/number of RBCs screened) * number of infected RBCs. Differences in sensitivity between miLab, and expert microscopy or RDT was calculated by McNemar’s test.

For Ethiopia, the presence of *P. falciparum* and *P. vivax* required special consideration for the analysis by expert microscopy. Because species can eventually be mixed up even by experienced microscopists, PCR correction of expert microscopy results was conducted as follows: (i) If expert microscopy identified *P. vivax*, but qPCR detected *P. falciparum* mono-infection, the result was corrected to *P. falciparum* (n=4). (ii) If expert microscopy identified *P. falciparum*, but qPCR detected *P. vivax* mono-infection, the result was corrected to *P. vivax* (n=9). (iii) If expert microscopy identified mixed infection with *P. falciparum* and *P. vivax*, but qPCR detected only *P. vivax*, the result was corrected to *P. vivax* (n=129). (iv) If expert microscopy identified mixed infection with *P. falciparum* and *P. vivax*, but qPCR detected only *P. falciparum*, the result was corrected to *P. falciparum* (n=2). No corrections were made if qPCR detected a mixed infection, but only one species was detected by expert microscopy (n=119). The reason for the large number of samples diagnosed as mixed infection by expert microscopy but as *P. vivax* mono-infection by qPCR is not known. The PCR result was largely confirmed by RDT; 122/129 samples were positive for *P. vivax* only, 1 for *P. falciparum* only, 4 were mixed species infections, and 2 were RDT-negative.

Likewise, the parasite species might be incorrectly identified by miLab. The miLab provides diagnosis as ‘suspected *P. falciparum*’, ‘suspected *P. vivax*’, ‘suspected *Plasmodium*’ (i.e., parasites were detected but the species was not identified), or ‘negative’. In case of any positive result, infected red blood cells are displayed on the screen of the device and the result can be verified by the operator. For the current study, initially sensitivity was calculated as the ability to detect any parasites, irrespective of whether the correct species was diagnosed. The number of samples diagnosed positive by miLab, but with the incorrect species identified, is presented separately. In line with data analysis for the miLab, microscopy conducted at the health center was counted as positive irrespective of species diagnosed, with the number of infections with the wrong species diagnosed presented separately.

### Sample size

This is a descriptive study. The sample size was determined to collect at least 200 *P. falciparum* and 200 *P. vivax* infections at densities >20 parasites/µL. The study was not powered to determine differences in sensitivity and specificity among diagnostic tools, e.g., miLab, RDT, and microscopy conducted at the health center. All analyses of comparisons that are reported are exploratory.

## Results

Across three sites in Ethiopia, and Ghana, a total of 2201 samples were collected and screened by miLab, RDT, expert microscopy, and qPCR. In Hawassa, Ethiopia, 1199 samples were collected and screened for *P. falciparum* and *P. vivax*. In Gondar, Ethiopia, 397 samples were collected and screened for *P. falciparum* and *P. vivax*. In Ghana, 605 samples were collected and screened for *P. falciparum*.

### Sensitivity and specificity of miLab

Across all sites, 28.9% (635/2201) of individuals tested positive by qPCR for *P. falciparum*. Sensitivity of miLab, RDT, and health center microscopy using expert microscopy and qPCR as gold standards are shown in Table 1. Using qPCR-positive infections at densities >200 parasites/µL as gold standard, miLab reached a sensitivity for *P. falciparum* of 97.4% (298/306) across all data, and of 94.2% to 100% across the three sites. Using a threshold of >20 parasites/µL by qPCR, the sensitivity of miLab was 91.8% (345/376) across all sites, and 78.2% to 100% per site. Using expert microscopy as reference, the sensitivity of miLab for *P. falciparum* across all sites was 96.3% (335/348).

**Table 1:** Sensitivity and specificity of miLab, microscopy conducted at the health center, and RDT using qPCR or expert microscopy as gold standard. For qPCR, sensitivity was calculated at thresholds of 20 and 200 parasites/µL. Specificity was calculated only including samples negative by qPCR for both species. For miLab, a test was considered positive if any parasites were detected, irrespective of species detected. As a result, specificity for miLab is identical for *P. falciparum* and *P. vivax*. Specificity using expert microscopy as gold standard was not calculated, as miLab and RDT were more sensitive than expert microscopy.

|  | Expert microscopy<br>as gold standard<br>[95% CI] (n/N) | qPCR as gold<br>standard [95% CI]<br>(n/N) |  |  |
| --- | --- | --- | --- | --- |
|  |  | Sensitivity at >200<br>parasites/uL | Sensitivity at >20<br>parasites/uL | Specificity |
|  | Sensitivity |  |  |  |
|  | miLab |  | miLab |  |
| ***P. falciparum*** |  |  |  |  |
|  | 97.5% [94.6, 99.1] | 97.8% [94.9, 99.3] | 93.8% [90.1, 96.4] | 98.0% [96.4, 99.1] |
| Hawassa | (232/238) | (219/224) | (242/258) | (500/510) |
|  | 98.1% [89.7, 100.0] | 100% [88.4, 100.0] | 100% [92.7, 100.0] | 96.7% [91.8, 99.1] |
| Gondar | (51/52) | (30/30) | (49/49) | (117/121) |
|  | 89.7% [78.8, 96.1] | 94.2% [84.1, 98.8] | 78.3% [66.7, 87.3] | 99.3% [98.0, 99.9] |
| Ghana | (52/58) | (49/52) | (54/69) | (436/439) |
|  | 96.3% [93.7, 98.0] | 97.4% [94.9, 98.9] | 91.8% [8.5, 94.3] | 98.8% [97.9, 99.4] |
| Combined | (335/348) | (298/306) | (345/376) | (1057/1070) |
| ***P. vivax*** |  |  |  |  |
|  | 95.7% [92.4, 97.8] | 95.9% [91.8, 97.4] | 88.5% [84.4, 91.8] | 98.0% [96.4, 99.1] |
| Hawassa | (243/254) | (256/267) | (277/313) | (500/510) |
|  | 98.7% [95.5, 99.8] | 95.9% [91.7, 98.3] | 89.9% [84.7, 93.8] | 96.7% [91.8, 99.1] |
| Gondar | (156/158) | (163/170) | (170/189) | (117/121) |
|  | 96.8% [94.7, 98.3] | 95.9% [93.6, 97.5] | 89.0% [86.0, 91.6] | 97.8% [96.3, 98.9] |
| Combined | (399/412) | (419/437) | (447/502) | (617/631) |
|  | **Health center** |  | **Health center** |  |
|  | **microscopy** |  | **microscopy** |  |
| ***P. falciparum*** |  |  |  |  |
|  | 91.2% [86.8, 94.5] | 83.5% [78.0, 88.1] | 76.0% [70.3, 81.1] | 99.8% [98.9, 100.0] |
| Hawassa | (217/238) | (187/224) | (196/258) | (509/510) |
|  | 67.3% [52.9, 79.7] | 50% [31.2, 68.7] | 32.7% [19.9, 47.5] | 99.2% [95.5, 100.0] |
| Gondar | (35/52) | (15/30) | (16/49) | (120/121) |
|  | 69.0% [55.5, 80.5] | 76.9% [63.2, 87.5] | 60.9% [48.4, 72.4] | 98.4% [96.7, 99.4] |
| Ghana | (40/58) | (40/52) | (42/69) | (432/439) |
|  | 83.9% [79.6, 87.6] | 79.1% [74.1, 83.5] | 67.5% [62.6, 72.3] | 99.2% [98.4, 99.6] |
| Combined | (292/348) | (242/306) | (254/376) | (1061/1070) |
| ***P. vivax*** |  |  |  |  |
|  | 90.6% [86.3, 93.9] | 71.9% [66.1, 77.2] | 62.3% [56.7, 67.7] | 99.2% [98.0, 99.8] |
| Hawassa | (230/254) | (192/267) | (195/313) | (506/510) |
|  | 77.2% [69.9, 83.5] | 72.4% [65.0, 78.9] | 66.7% [59.5, 73.3] | 99.2% [95.5, 100.0] |
| Gondar | (122/158) | (123/170) | (126/189) | (120/121) |
|  | 85.4% [81.7, 88.7] | 72.1% [67.6, 76.2] | 63.9% [59.6, 68.2] | 99.2% [98.2, 9.7] |
| Combined | (352/412) | (315/437) | (321/502) | (626/631) |
|  | **RDT** |  | **RDT** |  |
| ***P. falciparum*** |  |  |  |  |
|  | 92.9% [88.8, 95.8] | 97.3% [94.3, 99.0] | 93.4% [89.7, 96.1] | 99.0% [97.7, 99.7] |
| Hawassa | (221/238) | (218/224) | (241/258) | (505/510) |
| Gondar | 84.6% [71.9, 93.1] | 96.7% [82.8, 99.9] | 93.9% [83.1, 98.7] | 100% [97.0, 100.0] |
|  | (44/52) | (29/30) | (46/49) | (121/121) |
|  | 94.8% [85.6, 98.9] | 96.2% [86.8, 99.5] | 89.0% [76.7, 93.9] | 98.0% [96.1, 99.1] |
| Ghana | (55/58) | (50/52) | (60/69) | (430/439) |
|  | 92.0% [88.6, 94.6] | 97.1% [94.5, 98.6] | 92.3% [89.1, 94.8] | 98.7% [97.8, 99.3] |
| Combined | (320/348) | (297/306) | (347/376) | (1056/1070) |
| ***P. vivax*** |  |  |  |  |
|  | 98.1% [94.6, 99.6] | 89.9% [85.6, 93.2] | 78.0% [72.9, 82.4] | 99.0% [97.7, 99.7] |
| Hawassa | (155/158) | (240/267) | (244/313) | (505/510) |
|  | 91.3% [87.2, 94.5] | 95.3% [90.9, 97.9] | 87.3% [81.7, 91.7] | 100% [97.0, 100.0] |
| Gondar | (232/254) | (162/170) | (165/189) | (121/121) |
|  | 93.9% [91.2, 96.0] | 92.0% [89.0, 94.4] | 81.5% [77.8, 84.8] | 99.2% [98.2, 99.7] |
| Combined | (387/412) | (402/437) | (409/502) | (626/631) |

Across the two sites in Ethiopia, 44.4% (707/1589) of individuals tested positive by qPCR for *P. vivax*. Using qPCR-positive infections at densities >200 parasites/µL as gold standard, miLab reached a sensitivity for *P. vivax* of 95.9% (419/427) across Hawassa and Gondar, and of 95.9% in either site. Using a threshold of >20 parasites/µL by qPCR, the sensitivity of miLab was 98.0% (447/502) across the two sites. Using expert microscopy as reference, the sensitivity of miLab for *P. vivax* across the two sites was 96.8% (399/412).

Specificity was calculated as the number of samples with no parasite detected by miLab (irrespective of species detected) divided by the number of samples negative by PCR for either species. Across all sites, specificity of the miLab was 98.8% (1057/1070), and 96.7% to 99.3% per site.

While miLab missed 10/343 *P. falciparum* infections and 9/405 *P. vivax* infections detected by expert microscopy and confirmed *P. falciparum* and/or *P. vivax* positive by qPCR (including mixed infections), it detected 47 infections that were negative by expert microscopy but qPCR positive. Overall, miLab thus detected significantly more PCR-confirmed infections than expert microscopy (miLab: 65.1% (736/1131), expert microscopy: 62.5% (707/1131), *P*=0.0004).

Among samples positive by expert microscopy, miLab detected slightly more infections than RDT, while for samples positive by qPCR at densities >200 or >20 parasites/µL, miLab and RDT detected very similar numbers (Table 1, miLab vs. expert microscopy: *P*=0.0071; miLab vs. >200 parasites/µL by qPCR: *P*=0.7389; miLab vs. >20 parasites/µL by qPCR: *P*=0.2752). Among qPCR positive *P. falciparum* mono-infections of any density, miLab and RDT detected virtually the same number of infections (miLab: 176/258, RDT: 177/258, *P*=0.7389). Mixed-species infections were excluded from this analysis as it cannot be determined whether the miLab detected RBCs infected with *P. falciparum* or *P. vivax*, and thus the sensitivity of miLab might be overestimated.

Among samples positive for *P. vivax* by expert microscopy, or with densities >200 or >20 parasites/µL by qPCR, miLab detected moderately more infections than RDT (Table 1, miLab vs. expert microscopy: *P*=0.0047; miLab vs. >200 parasites/µL by qPCR: *P*=0.0007; miLab vs. >20 parasites/µL by qPCR: *P*<0.0001). No significant difference in sensitivity between miLab and RDT was observed among qPCR positive *P. vivax* mono-infections of any density (miLab: 333/496, RDT: 331/496, *P*=0.5930).

### Ability to diagnose the correct parasite species

The ability of miLab and microscopy conducted at the health center to diagnose the correct species was assessed in the two sites in Ethiopia. Because the miLab in its current set up cannot detect mixed infections (either *P. falciparum*, *P. vivax*, or *Plasmodium* is displayed), mixed infections by qPCR were excluded from this analysis. Samples diagnosed as mixed infections by health center microscopy with only one species detected by qPCR were assigned to the species detected by qPCR for this analysis.

Agreement between qPCR and miLab, calculated as Cohen’s kappa, was almost perfect for samples where miLab detected a species (kappa=0.942 for infections with densities >20 parasites/µL, Table 2), while agreement between qPCR and health center microscopy was substantial (kappa=0.745, Table 2). Among 177 *P. falciparum* mono-infections in Ethiopia at a density of >20 parasites/µL by qPCR, miLab diagnosed 147 correctly, while 1 was diagnosed as *P. vivax*, for 19 the species was not determined, and 10 were negative. miLab thus diagnosed the correct species in 99.3% (147/148) of infections where a species was determined (i.e., excluding negative samples and samples where ‘suspected *Plasmodium*’ was diagnosed), and in 88.0% (147/167) of infections where any parasites were detected (Table 2). In contrast, microscopy conducted at the health center diagnosed *P. falciparum* in 85.9% (128/149) of *P. falciparum* mono-infections by qPCR, while in 21/149 samples *P. vivax* was misdiagnosed (Table 2). Data for PCR-positive infections of all densities are given in Table 2.

**Table 2:**
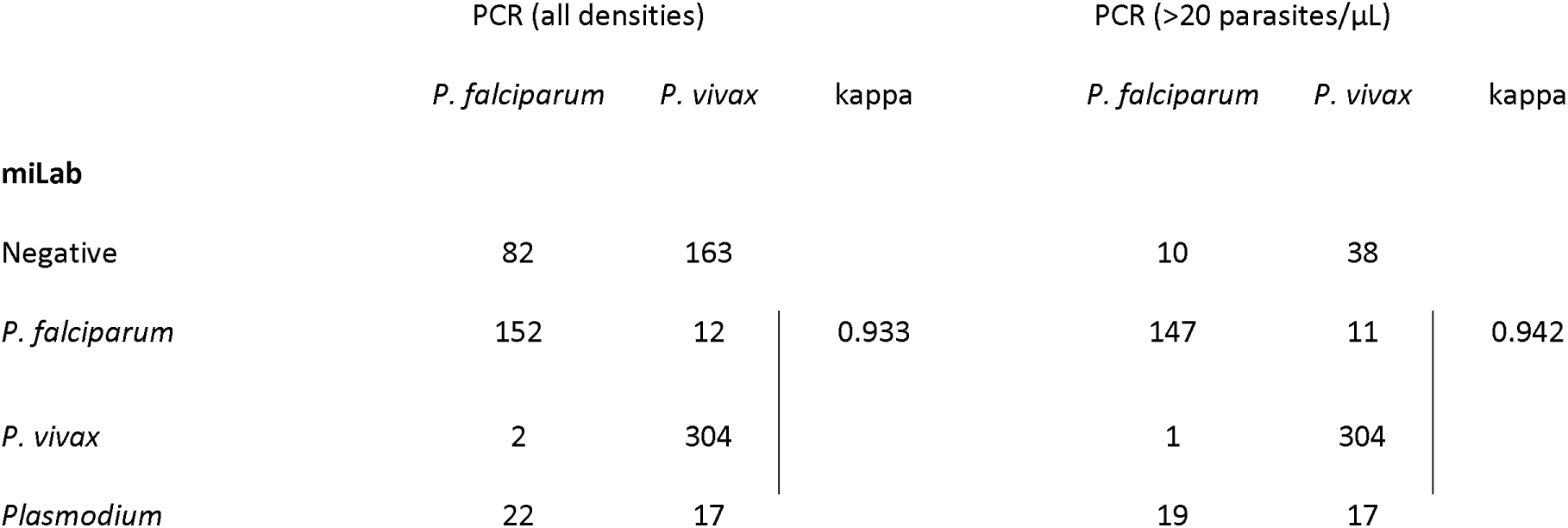
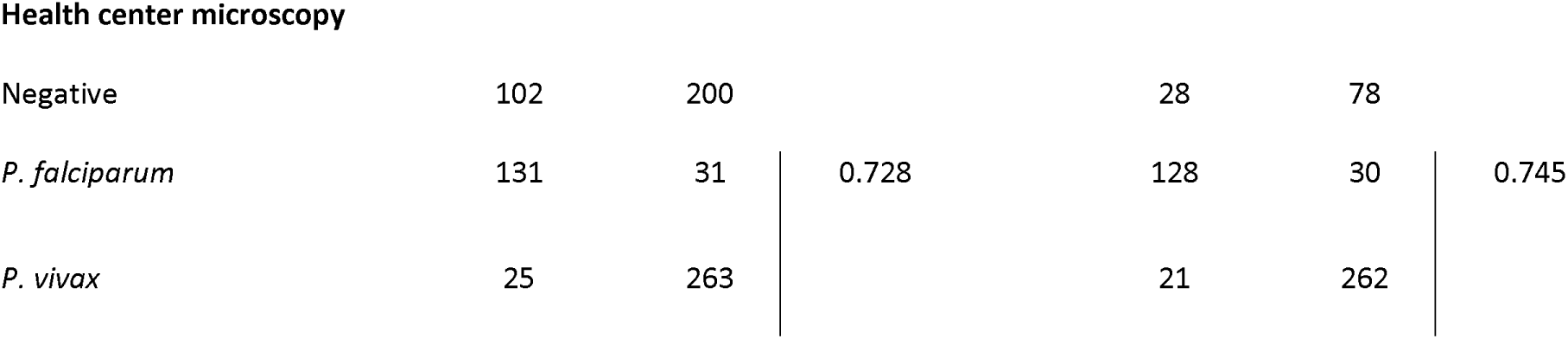
Ability of miLab and microscopy conducted at the health center in detecting the correct species among *P. falciparum* and *P. vivax* mono-infections by qPCR. To calculate kappa, samples negative by miLab or health center microscopy, and samples where miLab detected *Plasmodium* but did not determine the species, were excluded.

Among 370 *P. vivax* mono-infections at a density of >20 parasites/µL by qPCR, miLab diagnosed 304 correctly, while 11 were diagnosed as *P. falciparum*, for 17 the species was not determined, and 38 were negative (Table 2). miLab thus diagnosed the correct species in 96.5% (304/315) of infections where a species was determined, and in 91.6% (304/332) of infections where any parasites were detected (Table 2). Microscopy conducted at the health center diagnosed *P. vivax* in 89.7% (262/292) of *P. vivax* mono-infections by qPCR, while in 30/292 samples *P. falciparum* was misdiagnosed (Table 2). Data for PCR-positive infections of all densities are given in Table 2.

In 60 samples, miLab detected parasites but the species was not defined. By qPCR, 22/60 were *P. falciparum* mono-infections, 17/60 were *P. vivax* mono-infections, 17/60 were mixed infections, and 4/60 were negative. The number of infected red blood cells detected by miLab was lower in samples with no species determined compared to those where *P. falciparum* or *P. vivax* was suggested (*Plasmodium*: N=60, geometric mean=37.7; *P. falciparum*: N=314, geometric mean=178.8; *P. vivax*: N=371, geometric mean=223.3; Kruskal Wallis test *P*=0.0001). The small number of infections where an incorrect species was assigned by miLab prevented an analysis of parasite density distributions.

### Quantification of parasitemia

Density estimates between miLab and expert microscopy were assessed for *P. falciparum* and *P. vivax* mono-infections. Using log-transformed data, strong correlation was found for *P. falciparum* (Pearson’s r=0.72, *P*<0.001, Figure 1A) and moderate correlation *P. vivax* (Pearson’s r=0.54, *P*<0.001, Figure 1B). The ratio of the densities by miLab divided expert microscopy vs. density by expert microscopy was plotted. miLab tended to underestimate parasite density, and the underestimation grew as parasite density increased (Figures 1C, D). Similar trends were observed if density estimates by miLab were compared to those by qPCR (Supplementary Figure S1).

**Figure 1:**
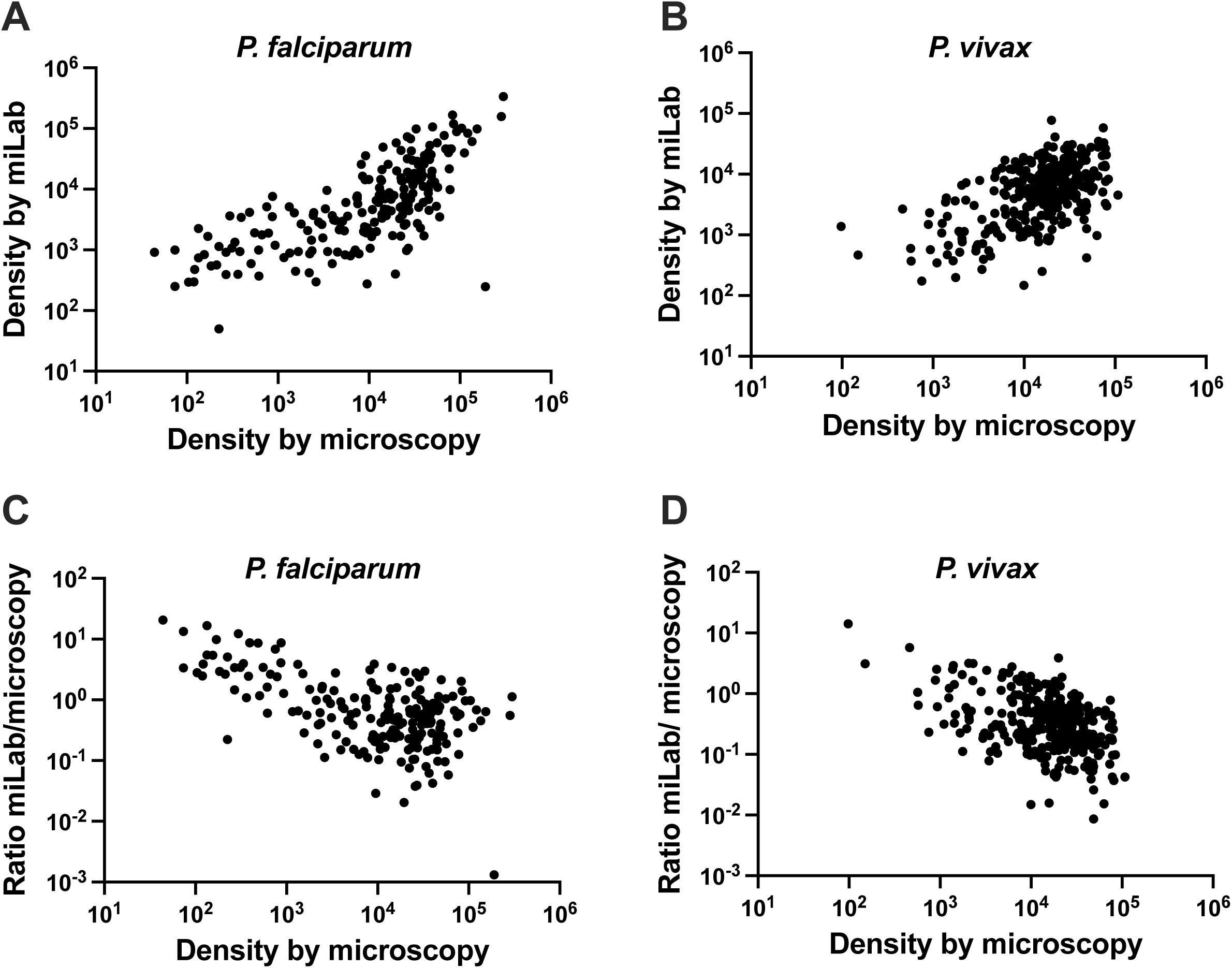
Correlation between quantification by expert microscopy and miLab for *P. falciparum* (A) and *P. vivax* (B). C, D: Ratio of densities by miLab divided by density by expert microscopy vs. the density by expert microscopy. Only single-species infections are included in all plots.

## Discussion

Automated microscopy for malaria diagnosis has the potential to overcome limitations of manual microscopy. The miLab automated microscope showed high sensitivity and specificity for *P. falciparum* and *P. vivax* diagnosis. At densities of over 200 parasites/µL, the miLab 97% of *P. falciparum* and 96% of *P. vivax* infections. The miLab detected more infections than microscopy conducted at the health center, and a similar number as the highly sensitive RDT used in this study. miLab also detected more PCR-confirmed infections than expert microscopy. In Ethiopia, where *P. falciparum* and *P. vivax* are co-endemic and correct diagnosis is a challenge, miLab assigned the correct parasite species to over 95% of infections detected.

The miLab showed clear advantages over microscopy conducted at health centers. In all sites, it detected more infections, and in Ethiopia assigned the correct species more often. Of note, in the current study, the sensitivity of the microscopists at the health centers and their ability to distinguish between *P. falciparum* and *P. vivax* was relatively high. Using expert microscopy as gold standard, microscopists at health centers detected parasites in 82.1% (611/717) of cases. In 10-14% slides diagnosed as positive the incorrect species was identified. This was better than observed in a nationwide comparison of expert microscopy and health center microscopy in Ethiopia, where parasites were detected on 70% of positive slides and the wrong species was assigned to almost one quarter of slides [4]. In other studies, as few as 50-60% of RDT-positive infections were detected by microscopists [24]. It cannot be ruled out that for the current study, microscopists at health centers spent extra time reading slides as they knew the data was recorded. Thus, under real world conditions, the benefit of miLab over microscopy conducted at the health center might exceed the effect observed here.

While the accuracy of the miLab to distinguish between *P. falciparum* and *P. vivax* was high, its inability to directly report mixed infections is a limitation. In Ethiopia, specific guidelines are in place for the treatment of mixed infections, namely artemether–lumefantrine plus primaquine to clear liver hypnozoites [25]. The miLab displays detected images of both *P. falciparum* and *P. vivax* on the result screen, allowing the user to potentially identify mixed infections. Additional studies are needed to evaluate the practical utility and effectiveness of this feature.

The miLab tended to underestimate parasite densities, in particular when parasite density was high. In many cases, the density by miLab was less than 10% of the density determined by expert microscopy or qPCR. An update to the algorithm that aims to improve parasite quantification is currently being evaluated. Once improvements are implemented, the ability of the miLab to determine densities can support treatment decisions, as very high parasitemia is a sign of severe malaria [26]. Quantification by miLab might also open up the possibility to use is for studies on drug resistance. Direct assessment of clinical resistance to the current first-line drug for *P. falciparum*, artemisinin, requires measurements of clearance half-life in patients or ex vivo [27, 28]. Parasite quantification for such studies can be challenging. Side-by-side comparisons using miLab and conventional methods for parasite density determination will be required to determine whether the integration of the miLab would be beneficial for drug resistance surveillance.

In conclusion, the miLab showed high sensitivity and specificity to diagnose malaria infections, and high accuracy to distinguish between *P. falciparum* and *P. vivax*.

## Supporting information

Suplementary File: database

Suplementary File: STROBE checklist

## Data Availability

All data produced in the present work are contained in the manuscript (Supplementary data base S1)

## Acknowledgements

The authors thank all study participants. They also thank personnel at health centers in Hawassa, Ethiopia, Gondar, Ethiopia, and Kumasi, Ghana. They recognize the contributions of Joeakim Baba Domosie from Ejisu Government Hospital as well as Dr. Veronica Bannor and Prince Nti from Asokwa Children Hospital in Kumasi.

## Supplementary Files

Supplementary data base S1 contains all data from the study.

Supplementary Figure S1: Correlation between quantification by qPCR and miLab for *P. falciparum*

Supplementary File: STARD checklist

