## Supplementary material for "AI-supported automated microscopy for malaria diagnosis": Suplementary Figure S1

Supplementary Figure

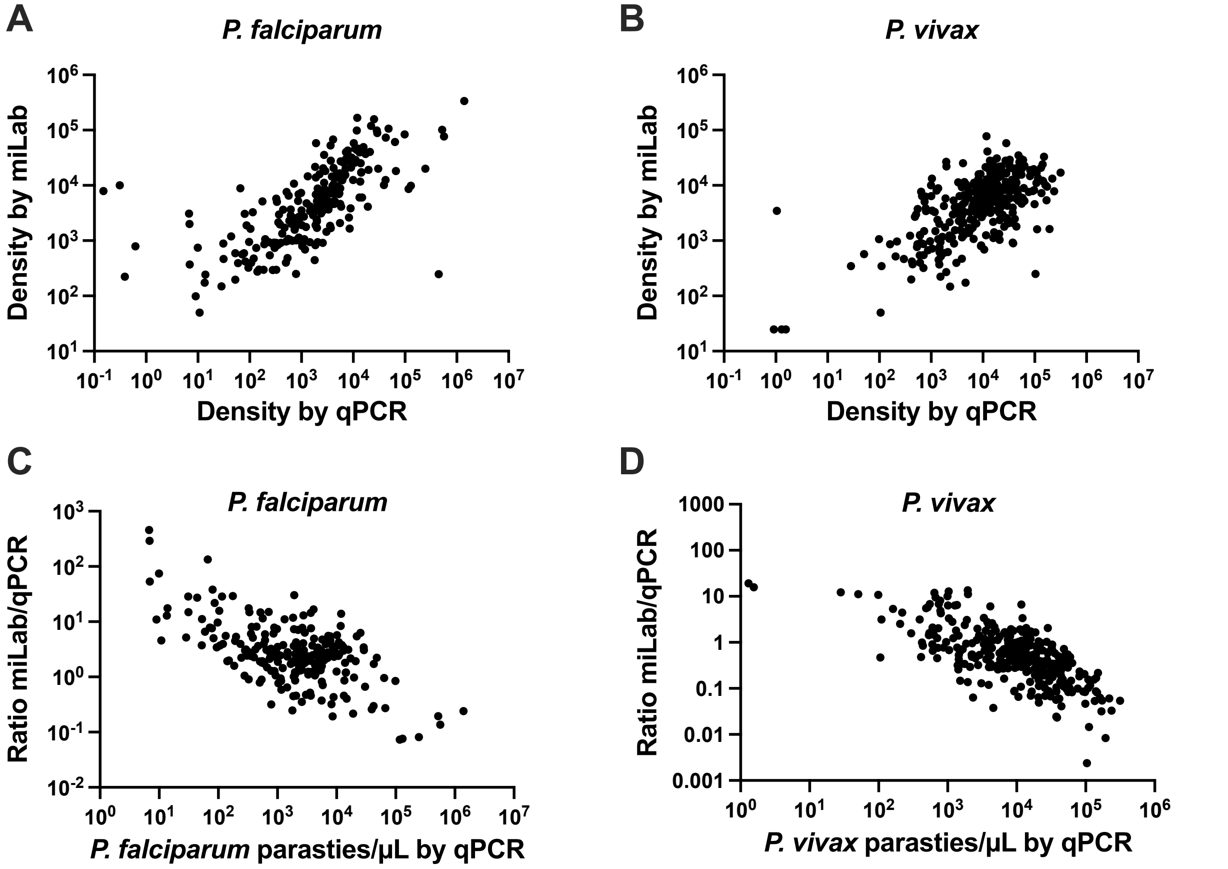


Figure S1: Correlation between quantification by qPCR and miLab for *P. falciparum* (A) and *P. vivax* (B). C, D: Ratio of densities by miLab divided by density by qPCR vs. the density by qPCR. Only single-species infections are included in all plots.
